# Impact of vaccination and risk factors on COVID-19 mortality amid delta wave in Libya: a single centre cohort study

**DOI:** 10.1101/2023.01.24.23284885

**Authors:** Inas Alhudiri, Zakarya Abusrewil, Omran Dakhil, Mosab Ali Zwaik, Mohammed Ammar Awn, Mwada Jallul, Aimen Ibrahim Ahmed, Rasha Abughrara, Adam Elzaghied

## Abstract

**Background:** The Delta variant has led to a surge in COVID-19 cases in Libya, making it crucial to investigate the impact of vaccination on mortality rates among hospitalized patients and critically ill.

**Aims:** To study risk factors and COVID-19 mortality rates among unvaccinated and vaccinated adults during delta wave at a single COVID-19 care centre in Tripoli, Libya.

**Methods:** The study involved two independent cohorts (n=341). One cohort was collected retrospectively from May 2021-August 2021 and the second cohort was prospectively collected from August 2021-October 2021 and most of them were during the Delta wave. The two cohorts were merged and analysed as one group.

**Results:** Most patients were male (60.5%) and 53.3% were >60 years. The vast majority of admitted patients did not have previous COVID-19 infection (98.9%) and were unvaccinated (90.3%). Among vaccinated, 30 patients had one dose and only 3 had two doses. Among patients who received one dose, 58.1% (18/31) died and 41.9% (13/31) survived. Most patients (72.2%) had a pre-existing medical condition. Multivariable prediction model showed that age >60 years was significantly associated with death (odds ratio=2.328, CI 1.456-3.724, p-value=<0.0001).

**Conclusion:** Previous infection or full vaccination against COVID-19 significantly reduces hospitalization and death, as most admitted patients were unvaccinated and not previously infected. However, a single vaccine dose may not be adequate, especially for older individuals and those with underlying medical conditions. High-risk older patients with comorbidities should be fully vaccinated and offered up to date bivalent COVID-19 booster doses.

## Introduction

The novel coronavirus (SARS-CoV-2) emerged in December 2019 and then spread around the globe. In Libya, the first case was identified on 24^th^ March 2020 by National Centre for Diseases Control (NCDC) in Tripoli in a pilgrim who came from Saudi Arabia; thereafter the cases continued to rise (1,2).

According to the reports from National Center for Diseases Control (NCDC), and until the time of writing this manuscript, the number of confirmed COVID-19 cases was 502,076, the number of vaccinated people with the first dose was 2,260,427, 1,177,469 with the second dose, and 117,799 with the booster dose (2).

COVID-19 vaccinations have been approved for emergency use worldwide (3). The humoral immune responses elicited by four vaccines used in Libya were previously evaluated in the general population (4). This study showed that Sputnik V and AstraZeneca vaccines developed a robust antibody response especially in previously infected individuals higher than Sinovac and Sinopharm.

Vaccination with either one dose of Pfizer BioNtech or AstraZeneca (Vaxzevria) was associated with a substantial reduction in symptomatic infection in adults older than 70 years and with additional protection against severe disease and hospitalization (5). Patients who had received one dose of AstraZeneca had an extra reduction of admission with severe disease (4). Furthermore, vaccination was associated with lower hospital mortality (6).

The onset of this pandemic has overextended the health systems of many countries, particularly the already limited intensive care space which is hugely required to care for those with severe illness (7). Therefore, understanding the risk factors and immunopathology of COVID-19 would help the clinicians prioritize critically ill patients and prevent the progression of the disease since increased admission to the intensive care unit (IUC) put the burden on the health care system which is associated with an increase in mortality rate. The infection fatality rate (IFR) and risk of death could be variable for each country due to health system variations, such as infrastructure, medical centres bed capacity, medical staff and availability of medicines (8).

The main factors associated with hospital admission were elderly age, male gender, ethnicity, fever, dyspnoea, comorbid diseases, cancer, occupation of healthcare workers, smoking, and pregnancy (9). Signs and symptoms associated with the need for ICU care were dyspnea, tachypnea and hypoxia (10).

Approximately 14% to 29% of hospitalized COVID-19 patients required intensive care due to acute respiratory distress syndrome (ARDS) where the mortality rate ranged from 8.7% to 21% among those patients with pneumonia (11–14).

The case fatality rate, defined as the ratio between confirmed deaths and confirmed cases, was between 1.7% - 1.4% in Libya during the study period (May-October 2021) (15). However, published data on the clinical characteristics and risk factors of COVID-19 and vaccination status and impact on mortality among hospitalized patients and critically ill in Libya was limited. Therefore, the objective of this study was to study risk factors and COVID-19 mortality rates among unvaccinated and vaccinated adults during delta wave in Libya at a single COVID-19 care centre in Tripoli, Libya.

## METHODS

### Study cohort and design

The study was conducted at Souq Thullatha Isolation Centre, a recently established COVID-19 dedicated healthcare isolation centre in Tripoli, Libya. As the epidemic evolved, the ministry of health has built new COVID-19 care facilities in Tripoli named as isolation centres with intensive care unit (ICU) beds, inpatient care and patient under investigation (PUI) units. Souq Thullatha isolation centre has 20 ICU beds, 27 inpatient capacity (ward) and 18 PUI. The study involved two independent cohorts. One cohort was collected retrospectively from May 2021-August 2021 and the second cohort was prospectively collected from August 2021-October 2021. The data from the two cohorts were independent, they were exposed to the same factors, they were in the same year and most of them were during the Delta wave. Hence the two cohorts were merged and analysed as one group. So this is a combined retrospective-prospective cohort study where all individuals admitted to the Souq Thullatha isolation centre over a period of 6 months from May 2021-October 2021 were enrolled.

The centre only admits COVID-19 positive patients defined as a positive result on real time reverse transcriptase quantitative polymerase chain reaction (RT-qPCR) assay of nasopharyngeal swab specimens or positive rapid antigen test in symptomatic patients. RT-qPCR Testing was conducted at COVID-19 diagnostic laboratories of Libyan Biotechnology Research Centre, Tripoli, Libya.

Moderate to severe patients who meet at least one of the following criteria: (1) symptoms of respiratory distress with a respiratory rate 30 times/min; (2) resting blood oxygen saturation less than 94%; (3) the imaging examination of lungs showing CORADS5 score.

We included in this study all patients admitted to the intensive care unit, and ward patients in the isolation center.

### Data collection and questionnaire design

Clinical data were collected from patients’ records according to a designated questionnaire and were entered into an anonymised database. The questionnaire included personal details, and admission detail of symptoms and its onset, treatment received, whether on oxygen mask, CPAP, or endotracheal intubation. It was also noted if the patient required renal dialysis, hemofiltration or psychiatrist intervention and if they developed cardiac or other complications and blood, plasma or immunoglobulin transfusion. Information on past medical history and pre-existing illnesses, vaccination details, date of vaccination, number of doses and any side effects; previous COVID-19 infection and its severity and number of reinfections were documented. The primary outcome was in-hospital mortality and the secondary outcome was development of severe complications. Full recovery was defined as patients who were either discharged or were still hospitalised but not ventilated at the end of the study.

### Standard treatment protocol

ICU patients with abnormal SpO2 and high CRP were started on IV dexamethasone (8 mg) until SpO2 >94. Patients were treated with doxycycline tablets 200 mg daily for 10 days, vitamin C 1000mg daily, zinc tablets 60mg daily and vitamin D3 2000 IU daily. In addition, Antipyretic (Paracetamol 1g) and proton pump inhibitors (famotidine 40 mg or pantoprazole 40 mg) were administered to most patients. Critically ill patients were given Intravenous (IV) broad-spectrum antibiotics.

Supplementary high flow oxygen was administered to achieve SpO2 of ≥94% using nasal prong or face masks (simple or non-rebreathing). Patients were placed on CPAP if they couldn’t reach target SpO2 with a 15L non-rebreather, and intubation and mechanical ventilation were for patients whose saturations continued to decline despite non-invasive ventilation.

Antiviral therapy (Remdisvir 100mg or Faviperavir 200 mg), immunosuppressants (Tocilizumab i.v/ Baricitinib 4 mg tablets), ipratropium bromide or ***N***-acetylcysteine nebulizer and anticoagulants were administered to selected patients according to their clinical presentation.

### Statistical analysis

All statistical analyses were conducted using SPSS v24 software. We used descriptive statistics to characterize each cohort of patients and categorical variables were compared in relation to 5 age groups (20 – 35, 35 – 50, 51-60, 61-70 and 71-105) using Chi square analysis.

We then fitted multivariable logistic regression models with death as the dependent variable to identify factors associated with those mortality. Two models were constructed for association with death. First, we constructed a model for all patient cohort including sex, age, vaccination status and comorbidity predictors (DM, hypertension, respiratory disease, cardiac disease, chronic kidney disease and neurological disease). Then, we constructed another model restricted to patients 60 years or younger using the same predictors. Univariate analysis was also performed for individual risk factors before they were added to the multivariable model. P-value less than 0.05 was considered to be statistically significant in all analyses.

## Results

### Characteristics of study population

During the study period, 349 patients were admitted to the isolation centre with COVID-19. Most patients were male (211 [60.5%]) with mean (SD) age of 61.7 (16.13) 61.7±16.14 years. Table 1 describes the clinical characteristics of the total cohort. Most patients (252 [72.2%]) had a pre-existing medical condition, including diabetes mellitus (163 patients [46.7%]), hypertension (142 patients [40.7%]), cardiac disease (44 [12.6%]), chronic kidney disease (34 patients [9.7%]), respiratory disease (23 [6.6%]), neurological disease (23 [6.6%]), liver disease (6 [1.7%]), Alzheimer (9 [2.6%]) and cancer (5 [1.4%]). The mean (SD) number of comorbidities was 1.4 (1.3) and 19.7% of patients had >2 comorbidities.

**Table 1.** Comparison of hospitalized patients during study period (May–October 2021) by vaccination status.

| **Characteristic** | **Vaccinated (%)** | **Unvaccinated (%)** | **P-value** |
| --- | --- | --- | --- |
| **N** | **34** | **315** |  |
| Sex (Male) | 28 (82.4) | 183 (58.1) | **0.006** |
| Age, years | 68.2±12.5 | 61±16.3 | **0.0129** |
| Pre-existing medical conditions | 27 (79.4) | 225 (71.4) | 0.324 |
| No of comorbidities |  |  |  |
| 0 | 15 (44.1) | 90 (28.6) |  |
| 1 | 11 (32.4) | 98 (31.1) |  |
| 2 | 4 (11.8) | 65 (20.6) |  |
| >2 | 4 (11.8) | 62 (19.7) |  |
| Hypertension | 16 (47.1) | 126 (40) | 0.426 |
| Diabetes | 19 (55.9) | 144 (45.7) | 0.259 |
| Cardiac disease | 7 (20.6) | 37 (11.7) | 0.140 |
| Chronic kidney disease | 3 (8.8) | 31 (9) | 0.849 |
| Respiratory disease | 1 (2.9) | 22 (7) | 0.367 |
| Neurological disease | 1 (2.9) | 22 (7) | 0.367 |
| Previous COVID-19 | 1 (2.9) | 3 (1) | 0.301 |
| Death | 18 (52.9) | 183 (58.1) | 0.56 |
| One dose | 31 (91.2) | N/A | - |
| Two doses | 3 (8.8) | N/A | - |

The Mean age of death was 64.4 years, mean age of death in patients with chronic disease was 65.7 years and the mean age of death in patients who didn’t have chronic disease was 59.6 years. The overall death rate was 57.7% (201/349), patients 60 years or less the death rate was 75/163 (46%) and those >60 years was 125/185 (67.6%) [Figure.1].

**Figure 1.**
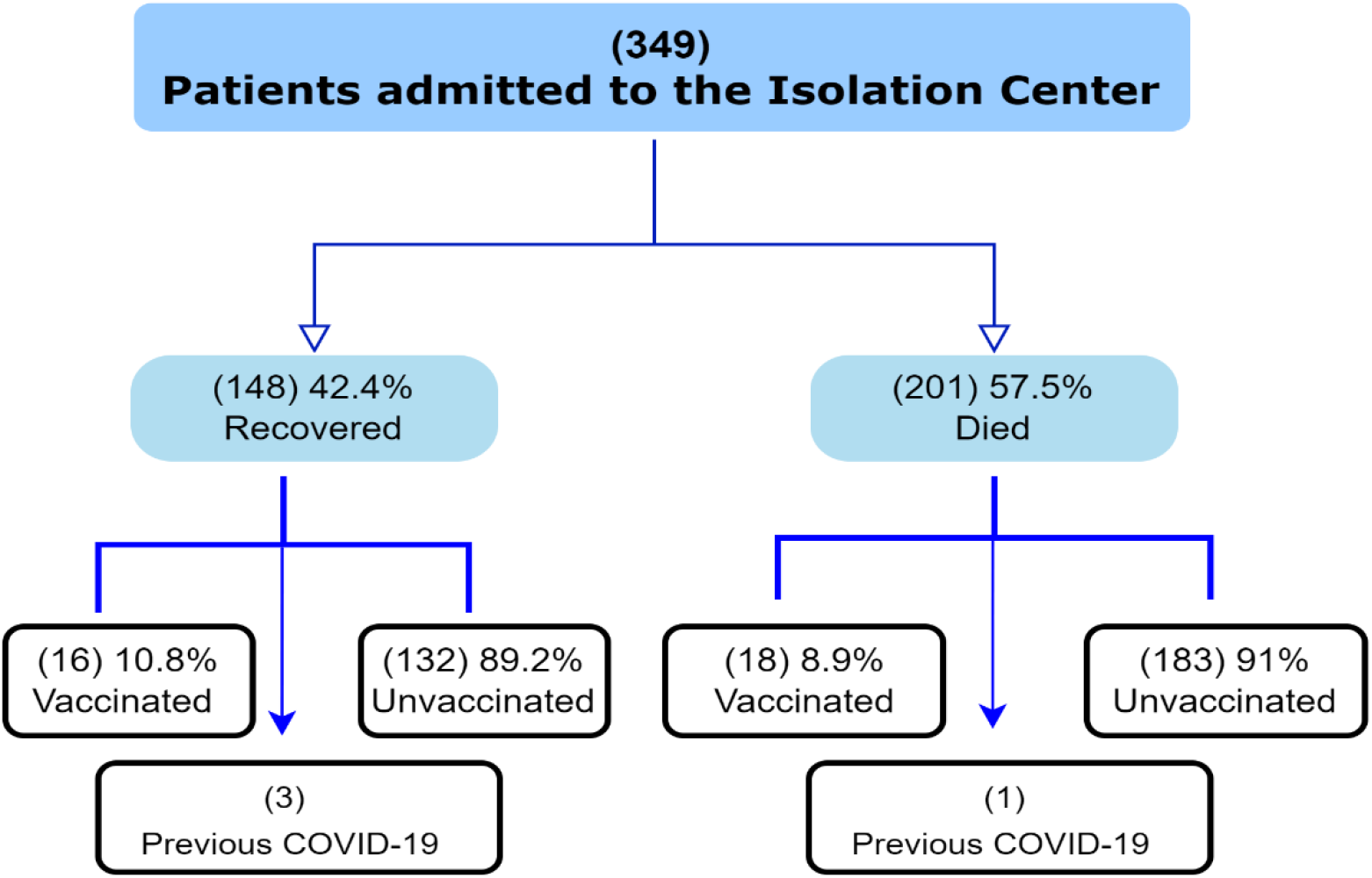
Outcome of study cohort.

### Comparison of vaccinated and unvaccinated COVID-19 patients

The vast majority of patients were not vaccinated against COVID-19 (315 [90.3%]) and were not previously infected (345/349 [98.9%]) (Table 1). The proportion of individuals who received one dose of the vaccine was 14.1% (30/213), while the proportion of individuals who received two doses of the vaccine was 1.4% (3/213). A comparison of vaccine coverage between genders revealed a statistically significant difference, with 13.3% (28/211) of male patients being vaccinated in contrast to only 4.3% (6/138) of female patients (p=0.006).

The 3 patients who received two doses had recovered and discharged from the hospital. Two patients got Sinopharm and AstraZeneca vaccines, while the third patient received an unknown type of vaccine. The three individuals, including one without pre-existing medical condition, were aged between 47 and 78 years old.

The mortality rate among patients who received one dose of the vaccine was 58.1% (18/31). A majority of the individuals who died were male (77.8%, 14/18) and had an age above 60 years (89%, 16/18). Furthermore, 77.8% (14/18) of the deceased had pre-existing medical conditions. Among patients with comorbidities, 50% (9/18) had hypertension, 55.6% (10/18) had diabetes, 16.7% (3/18) had cardiac disease, and 5.5% (1/18) had chronic kidney disease. The remaining four individuals (4/18) did not have any comorbidities, two of them received AstraZeneca vaccine, one had Sputnik V and the third had an unknown type of vaccine. These patients were aged between 58-85 years old. One of them received the vaccine just three days before hospital admission, while the other three received it 78-116 days before admission.

Of the patients who received one dose of the vaccine, one had a previous COVID-19 infection. This individual was male >65 years who died due to cardiac complications. His previous COVID-19 infection was mild, characterized by loss of smell and taste. The patient received one dose of the Sputnik V vaccine approximately 57 days prior to his illness. Among patients vaccinated with one dose who had comorbidity and died (9), 4 were vaccinated with AstraZeneca, 4 with Sputnik V, and 4 with Sinopharm. One patient didn’t report the vaccine type. The duration between vaccination and admission was less than 45 days in 7 patients, >45 days in 5 patients.

Among those vaccinated with one dose and had full recovery (13/31), the majority were men (11/13) with a mean age of 64.6 years. Four had received Sputnik V, 2 had AstraZeneca, 3 had Sinopharm and 4 had unknown vaccine type. About 85% (11/13) had comorbidity including hypertension, diabetes, chronic kidney disease and cardiac disease.

Also among patients with previous COVID-19 infection, 75% (3/4) had complete recovery and 25% (1/4) died from COVID-19 during hospital stay.

### Comparison of different periods of the COVID-19 pandemic

The mortality rates were calculated for each month as follows: 33.3% in May, 48.8% in June, 63.1% in July, 78.3% in August and 44.7% in September and October (p value≤0.0001) [Figure.2]. From mid-June to October, this period was predominantly Delta strain in Libya (16).

**Figure 2.**
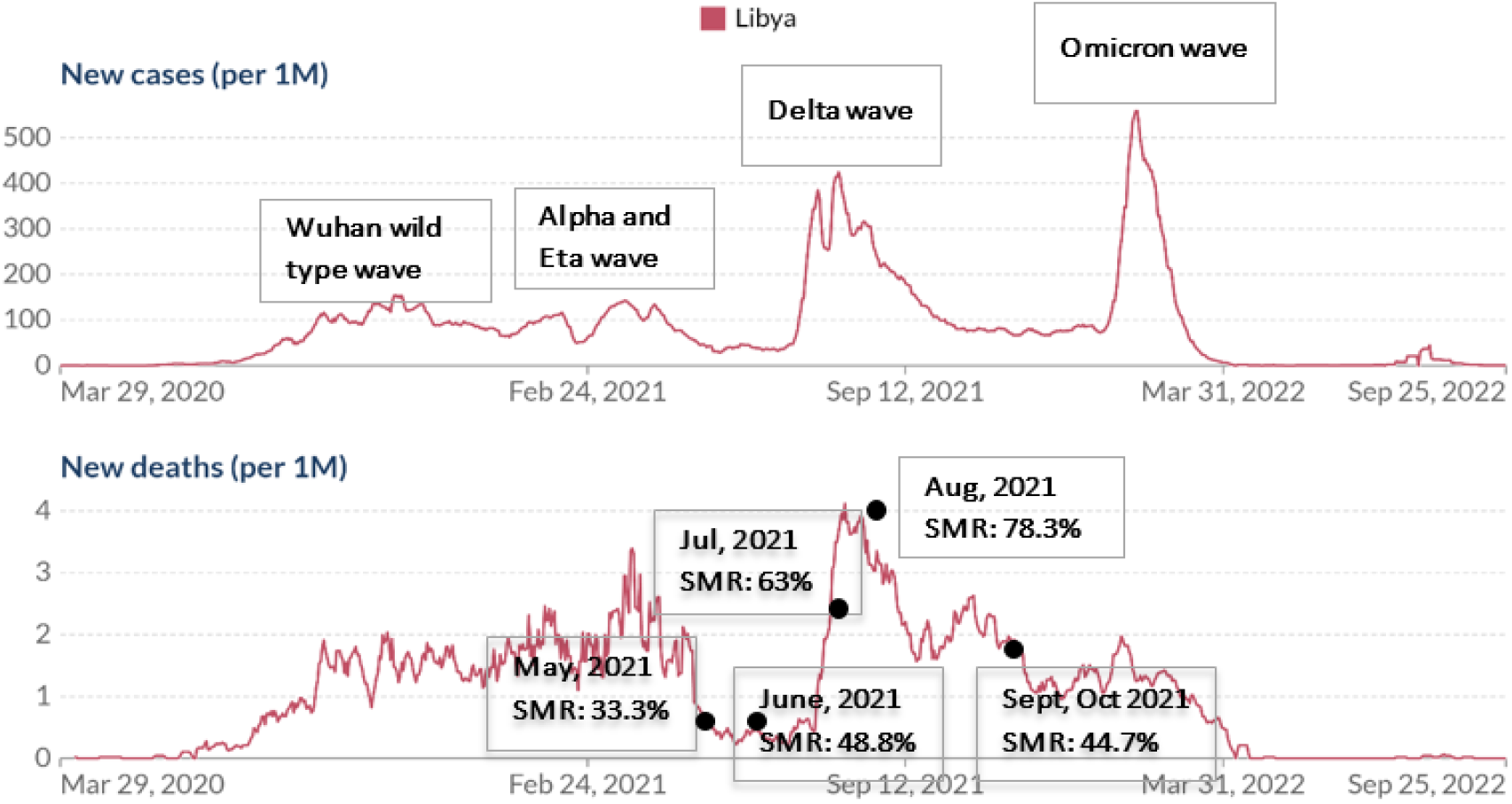
Seven-day rolling average of COVID-19 new confirmed cases in Libya and mortality rates in 2021 showing the different COVID-19 epidemic peaks and hospital mortality rates in this study. SMR= study mortality rate

### Predictors of mortality

In univariate analysis, factors most strongly associated with death were age >60 years (OR=2.46, 95% confidence interval 1.6-3.8, p-value =<0.0001) and hypertension (OR=0.6, 95% CI 0.39-0.94. p-value= 0.025).

In addition, univariate analysis showed that the mortality increases with increasing number of pre-existing medical conditions the patient have. The odds of death increases 2.8 fold in patients with >2 conditions (p-value 0.008) compared with those who doesn’t have chronic disease on presentation and 2 fold in those with at least 1 comorbid condition (p-value=0.010). We constructed a multivariable prediction model using sex, age, chronic diseases and vaccination status as possible risk factors for the patient cohort (Table 2). The model showed that the only significant factor was age older than 60 years (OR=2.328, CI 1.456-3.724, p-value=<0.0001).

**Table. 2.** Multivariable risk model for mortality.

| **Characteristic** | **Adjusted Odds ratio**<br>**(95% CI)** | ***P*-value** |
| --- | --- | --- |
| Sex (Female) | 1.550 (0.970-2.477) | 0.067 |
| Age >60 | 2.328 (1.456-3.724) | **<0.0001** |
| Hypertension | 0.789 (0.478-1.302) | 0.354 |
| Diabetes | 0.837 (0.528-1.326) | 0.448 |
| Chronic kidney disease | 0.991 (0.454-2.166) | 0.983 |
| Cardiac disease | 1.123 (0.551-2.288) | 0.750 |
| Respiratory disease | 0.655 (0.250-1.716) | 0.390 |
| Neurological disease | 0.787 (0.303-2.041) | 0.622 |
| Vaccination status | 1.450 (0.677-3.106) | 0.339 |

## Discussion

This study describes the outcome and clinical characteristics of 349 patients with COVID-19 admitted to Souq Thullatha center, a COVID-19 dedicated care center in Tripoli, Libya during the Delta wave in the context of vaccination status and other parameters.

Our study showed that 57.5% of patients has died due to complications of COVID-19 during their hospital stay. The mortality rate reported in other studies ranged from 8.7% to 54.6% among those patients with pneumonia (17–20). The high mortality rate and poor ICU outcomes led to concerns regarding the effectiveness of standard of care delivery and mechanical ventilation measures (21).

Most patients admitted were either not vaccinated (90%) nor previously infected with SARS-COV-2 (98.8%). This suggests that previous infection (natural induced immunity) and full vaccination against COVID-19 significantly reduces hospitalization and death. Furthermore, there was no mortality among fully vaccinated patients. Our cohort have only 3 fully vaccinated patients which suggests that two dose vaccine most probably protected individuals from severe disease warranting admission. At the time of conducting this study, the Libyan COVID-19 vaccination program has already started in April 2021 and by October 2021 about 1,393,030 persons had received one dose and 211278 had two doses according to the NCDC data. The total number of infected individuals in the NCDC registry was 341091 on 30^th^ September 2021. Our results thus confirm the findings of previous studies on the protective effect of full vaccination as single dose vaccine immunity tends to wane faster and may not mount a good immune response after infection except in previously infected individuals (4). We also found no difference in in mortality between unvaccinated and partially vaccinated. This is in agreement with other studies which additionally showed a significant difference in mortality between fully vaccinated and either unvaccinated or partially vaccinated similar to our findings (22). Our results indicate that whether a person has a history of infection or is only partially vaccinated, once admitted with severe disease, the risk of death is essentially the same as unvaccinated naïve individuals.

In addition, the mortality rates in this study were highest in July and August coinciding with delta predominance in most countries (23–25), then moderately decreased in September and October. This is probably due to increased vaccination coverage and provision of the second dose to people who had Sputnik V as their first dose. mRNA COVID-19 vaccination had also started in early August enhancing the vaccine resources in Libya (26).

In our study cohort, 72.2 % of patients had at least one pre-existing condition and about 20% had >2 comorbidities. Other studies showed similar findings and the proportion ranged between 65-80% of patients admitted to hospital had at least one of eight major chronic diseases (27,28). Our results found that a high percentage of patients were of advanced age with comorbidities, diabetes and hypertension being the most common and more likely to be male. These findings support the observations of other studies (10,18,29).

Studies showed that race, male sex, severe obesity and chronic kidney disease were significantly associated with the need for ICU care (10,28). Furthermore, previous studies showed that patients admitted with pre-existing disease especially heart failure and chronic kidney disease, old age and male have worse prognosis (27,30). However, in our study there was no statistically significant difference between male and female sex in the probability of death when sex was adjusted with other factors (table 2). When univariate analysis was carried out for each risk factor female patients were more likely to die than male patients. This finding was also seen when subgroup multivariate analysis was performed for patients <60 years. These differences could be explained by lower vaccination coverage in our female cohort (4.3% vs 13.3%). Individual traits including age, obesity, and pre-existing illness, are known to have an impact on immunological competence (31).

The vaccination campaign in Libya started on 17^th^ April 2021 with first dose of AstraZeneca and Sputnik V vaccination. On 26^th^ August 2021, 10, 356, 40 persons had first vaccine dose while 32693 had 2^nd^ dose. Unfortunately, many people who received Sputnik V first dose didn’t receive 2^nd^ dose until October-December and most of them received Astrazeneca as a second dose. In addition, Pfizer-BioNtech vaccination started later in the beginning of August. Our study period (mid-June to October) coincided with the emergence of the Delta SARS-COV-2 variant (16). SARS-COV-2 samples from Libya were regrettably not confirmed by genome sequencing during that time; instead, tracking relied on RT-qPCR assays that targeted the presence of the L452R and P681R mutations. However, samples collected around the end of February and beginning of March 2021 had their genomes sequenced, and the majority of cases were B.1.525, with only 2% being an alpha variant (32). Hence, we believe that most of SARS-COV-2 infection in May and beginning of June 2021 was B.1.525 variant.

Our study has a number of limitations. First, this study was an observational study conducted at a single center in Tripoli area thus the results might not reflect the situation in other Libyan COVID-19 isolation centres. Also, many patient medical records was missing COVID-19 vaccination type and doses. Additionally, although a multivariate regression analysis was performed to studying multiple factors associated with mortality, potential confounders may still become unrecognized. Our information regarding patient death is limited for hospital stay and patients were not followed-up after discharge. Nevertheless, our results provide important data on mortality rates and associated risk factors and inform public health COVID-19 control efforts. Further investigation should study other unmeasured factors including inflammatory markers to predict factors associated with mortality in COVID-19 patients admitted to the ICU. It is important to note that the political and economic instability has hugely affected the response to the pandemic, especially the provision of timely vaccine doses, oxygen supplies, capacity building for physicians and availability of antiviral and supportive therapy.

## Conclusion

Overall, we found that age and comorbidities were strong predictors of mortality. Clinicians should consider old patients with comorbidities as very high risk and treated with maximum care. High mortality rates raised concerns on the effectiveness of standard of care delivery and assisted ventilation measures. Health sector improvement plans should strongly consider capacity building in critical care and pandemic preparedness among the priority reform strategies.

The risk models created in this study may be used in a variety of health care settings, in the pandemic period to assess individual risk for death and guide patient management.

### Vaccination

Health recording systems in isolation centres should be improved and all patient clinical details should be registered, this could be achieved by strong vigilance and audit. Regular reporting to relevant authorities is suggested to quickly identify missing data files.

## Data Availability

All data produced in the present study are available upon reasonable request to the authors

## Acknowledgments

While it is not possible to honour all of the Libyan health workers who have died from COVID-19, the authors would like to pay special tribute to Dr. Firas Fadel who lost his life to the disease while serving his patients. The authors would like to thank the Libyan Ministry of Health and the Libyan Biotechnology Research Center for their support.

## Author contribution

IA, ZA, and OD conceptualised and designed the study. ZA, OD, MZ, MA, and AH prepared the study data. IA and RA performed the statistical analysis. All authors contributed to interpretation of the findings. IA, ZA and MJ wrote the original draft. All authors contributed to review and editing of the manuscript and approved the final manuscript. The corresponding author confirms that all authors meet authorship criteria and that no others meeting the criteria have been omitted.

## Ethical Approval

This study was conducted in accordance with the amended Declaration of Helsinki. The study was approved by the Bioethics Committee of the Libyan Biotechnology Research Center, Tripoli, Libya.

