## supplemental table 1 for "Impact of vaccination and risk factors on COVID-19 mortality amid delta wave in Libya: a single centre cohort study"

**Table 2. General characteristics of study cohort**

| Characteristic | Total | Age categories N (%) | | | | | P-value |
| --- | --- | --- | --- | --- | --- | --- | --- |
|  |  | 20 – 35 | 36 – 50 | 51-60 | 61-70 | 71-105 |  |
| Mean age ± SD |  | 30.15±3.95 | 44.8±4.4 | 55.8±3.04 | 65.9±2.7 | 79.8±7 |  |
| All Cohorts | 349 | 20 (5.7) | 77 (22) | 66 (18.9) | 74 (21.2) | 112 (32.1) |  |
| Sex |  |  |  |  |  |  |  |
| M | 211 | 16 (7.6) | 44 (20.8) | 47 (22.3) | 42 (20) | 62 (29.4) | 0.078 |
| F | 138 | 4 (2.9) | 33 (23.9) | 19 (13.8) | 32 (23.2) | 50 (36.2) |  |
| Previous COVID-19 diagnosis |  |  |  |  |  |  |  |
| Yes | 4 | 2 | 0 | 0 | 2 | 0 |  |
| No | 345 | 18 | 77 | 66 | 72 | 112 |  |
| Comorbidity |  |  |  |  |  |  |  |
| Yes | 252 | 7 (2.8) | 41 (16.3) | 50 (19.8) | 64 (25.4) | 90 (35.7) | <0.0001 |
| No | 97 | 13 (13.4) | 36 (37.1) | 16 (16.5) | 10 (10.3) | 22 (22.7) |  |
| No of comorbidities |  |  |  |  |  |  |  |
| 0 | 97 | 13 (13.4) | 36 (37.1) | 16 (16.5) | 10 (10.3) | 22 (22.7) |  |
| 1 | 111 | 3 (2.7) | 21 (18.9) | 23 (20.7) | 29 (26.1) | 35 (31.5) | <0.0001 |
| 2 | 72 | 1 (1.4) | 8 (11.1) | 18 (25) | 18 (25) | 27 (37.5) |  |
| >2 | 69 | 3 (4.3) | 12 (17.3) | 8 (11.6) | 17(24.6) | 28 (40.5) |  |
| Unvaccinated | 315 | 20 (6.3) | 74 (23.5) | 62 (19.7) | 62 (19.7) | 97 (30.8) | 0.059 |
| Any vaccine  One dose  Two doses | 31  3 | 0  0 | 2 (6.5)  1 (33.3) | 4 (12.9)  0 | 12 (38.7)  0 | 13 (41.9)  2 (66.7) |  |
| AstraZeneca  Sputnik V  Sinopharm  Unknown | 9  9  8  8 | 0  0  0  0 | 1 (11.1)  0  0  2 (28.6) | 0  1 (11.1)  0  3 (42.9) | 1 (11.1)  6 (66.7)  5 (62.5)  0 | 7 (77.8)  2 (22.2)  3 (37.5)  3 (37.5) |  |
